# Assessing Radiotherapy Responses in Low-Risk Prostate Cancer: A Longitudinal Analysis of T2 Relaxation Times

**DOI:** 10.1101/2023.07.01.23292072

**Authors:** Pavla Hanzlikova, Dominik Vilimek, Radana Vilimkova Kahankova, Martina Ladrova, Valeria Skopelidou, Zuzana Ruzickova, Radek Martinek, Jakub Cvek

## Abstract

**Rationale and Objectives:** To evaluate short and long-term changes in *T*_2_ relaxation as a response to radiotherapy in patients with low and intermediate risk localized prostate cancer.

**Materials and Methods:** A total of 24 patients were selected for this retrospective study. Each participant underwent 1.5T magnetic resonance imaging on seven separate occasions: initially after the implantation of gold fiducials, the required step for Cyberknife therapy guidance, followed by MRI scans two weeks post-therapy and monthly thereafter. As part of each MRI scan, the prostate region was manually delineated, and the *T*_2_ relaxation times were calculated for quantitative analysis. The *T*_2_ relaxation times between individual follow-ups were analyzed using Repeated Measures Analysis of Variance.

**Results:** Repeated Measures Analysis of Variance (RM-ANOVA) revealed a significant difference across all measurements (F (6, 120) = 0.611, p *<<* 0.001). A Bonferroni post hoc test revealed significant differences in median *T*_2_ values between the baseline and subsequent measurements, particularly between pre-therapy (*M*_0_) and two weeks post-therapy (*M*_1_), as well as during the monthly interval checks (*M*_2_ - *M*_6_). Some cases showed a delayed decrease in relaxation times, indicating the prolonged effects of therapy.

**Conclusion:** The changes in *T*_2_ values during the course of radiotherapy can help in monitoring radiotherapy response in unconfirmed patients, quantifying the scarring process, and recognizing the therapy failure.

## Introduction

Prostate cancer (PCa) is one of the most common malignant tumors in middle-aged and elderly men [1]. There is a clearly increasing incidence of this disease, which threatens not only the physical but also the psychological health of these patients [2]. Magnetic resonance imaging (MRI) plays a vital role in the PCa detection, as it enables to quantitatively objectify the state of the tissue. The standardly used screening methods using prostate-specific antigen (PSA) does not provide reliable information about the presence of PCa and there are many factors influencing the results [3]. Examinations using MRI reported higher sensitivity and specificity compared to other methods, such as positive prostate antigen blood test or transrectal ultrasound biopsy (TRUS) [4], which is, moreover, invasive and it is hard to decide the necessity of such examination to not harm a patient [3]. On the other hand, MR is a non-invasive technique providing high soft-tissue contrast and resolution, and is free of radiation compared to widely used computed tomography (CT) [3]. However, diagnosis using MRI is time consuming and demands a substantial expertise, as large number of 3D images needs to be read [4] and human image analysis is prone to errors in interpretation caused by observer limitations and complexity of clinical cases [5].

More accurate disease detection can be achieved by using multiparametric MRI (mpMRI) examinations. As the name implies, mpMRI is based on the use of multiple parameters, for example *T*_2_ weighting, diffusion-weighted image (DWI), and dynamic postcontrast examination (DCE - dynamic contrast enhancement) along with using Prostate Imaging and Data Reporting System (PI RADS) v2.1 [6]. The main function of the mpMRI is to quantitatively objectify the state of the tissue. An effective way how to objectively describe changes in the tissue behavior is relaxometry, which allows to reflect tissue properties and histological changes by the measurement of tissue relaxation times, such as *T*_1_, *T*_2_, and 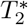, with the possibility to differentiate grades of PCa, especially in combination with mapping of Apparent diffusion coefficient (ADC). Previous studies showed that *T*_2_ and 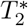 are shortened in more aggressive cancers compared to low-grade cancers [7]. The most common and easiest method is to perform *T*_2_ relaxometry, which uses transverse relaxation time description from different tissues [8]. Interpretation of the imaging data can be simplified by separating these independent sources by directly calculating the proton density and spin relaxation times. *T*_2_ map acquisition can facilitate improved tissue characterization, increase image tissue contrast, and provide a more direct link between observed signal changes and microanatomical ones. Furthermore, the quantitative nature of the data enables easy comparisons across longitudinal time points [9]. The benefit of *T*_2_ relaxometry is that it is a non-contrast technique, so could potentially decrease the risks and costs of contrast administration [7] and uses scanning at multiple echo times without the need for image normalization. These metrics describe the internal structure and heterogeneity within tumors and give more information about the disease than traditional whole-tumor assessments.

The aim of this paper is to investigate whether the MRI *T*_2_ mapping analysis can quantitatively represent PCAa intratumor heterogeneity and provide sensitive and robust insight into response to radiotherapy.

## Material and Methods

The purpose of this paper is to explore the *T*_2_ relaxation times in patients undergoing radiotherapy. The study provides an in-depth analysis of the changes in these times as well as their potential correlation with therapeutic outcomes. For a simplified overview of the study, please refer to Figure 1, which presents a diagrammatic representation of the study setup.

**Figure 1:**
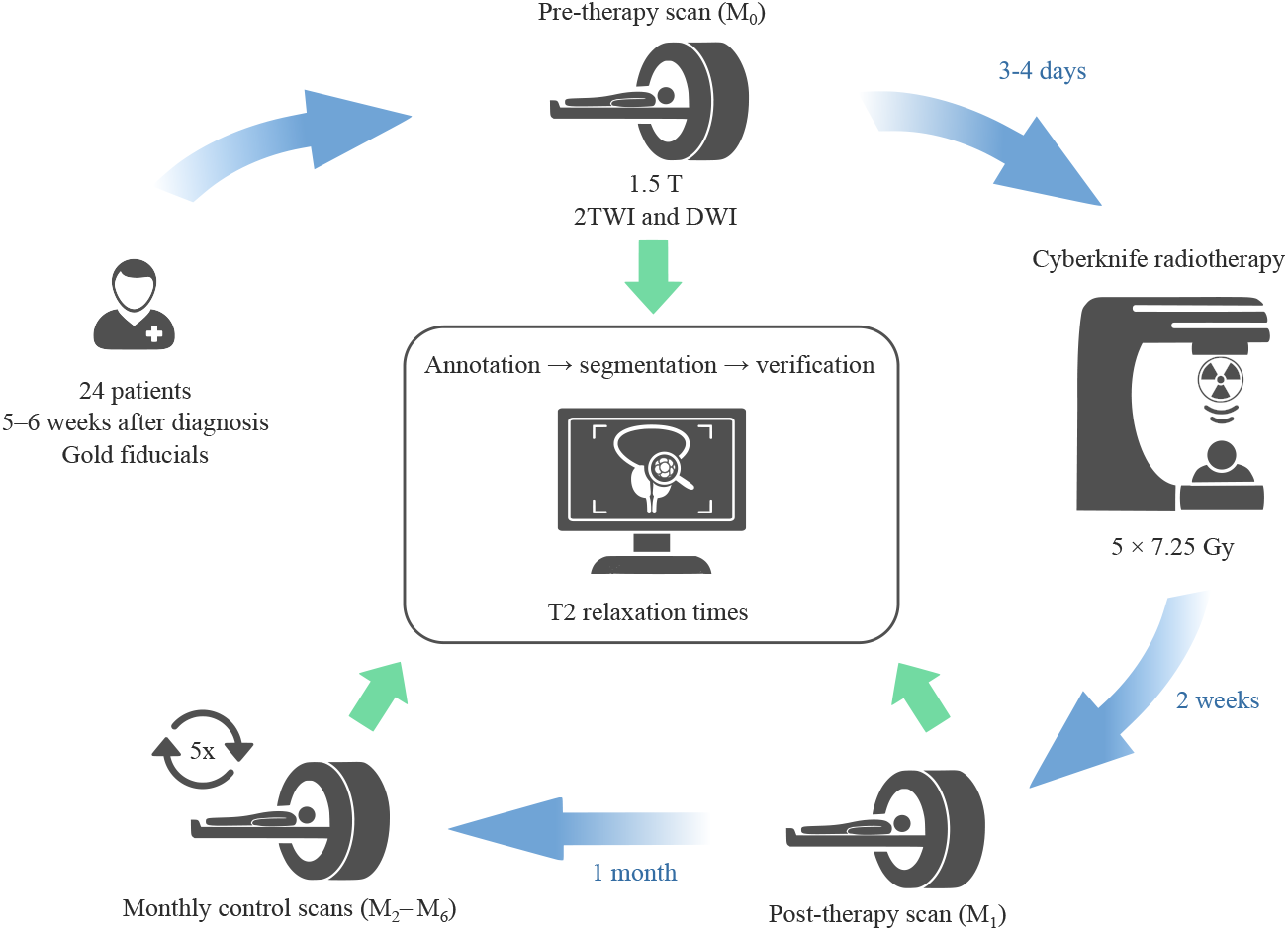
A simplified study workflow - A visual summary of the research design and methodology employed in the study, detailing patient selection, therapy application, MR examinations and MR data analysis.

### Patients Characteristics

A total of 24 patients (mean (standard deviation) age: 72.96 (6.29) years, age range: 61 *−*85 years) who underwent radiotherapy using Cyberknife for low-risk prostate cancer were selected (Gleason score 3 + 3 and 3 + 4, staging T1, N0 or T2aN0). All participants included in the study did not undergo any other treatment for prostate cancer and were patients of the Oncology Clinic of the University Hospital in 2020-2022. All patients underwent implantation of gold fiducials for navigation for the Cyberknife (fiducials are not a contraindication for MR imaging and radiotherapy according to studies [10, 11]). Patients with previous chemotherapy, radiotherapy or hormone replacement therapy were excluded. Informed consent form was obtained from all subjects and this study was approved by the institutional review board by University Hospital (RVO-FNOs/2020).

### Therapy

Treatment was started within 5-6 weeks from diagnosis. The gold fiducials were introduced for the navigation of robotic radiotherapy 3 or 4 days prior the actual irradiation. Radiation treatment was then carried out in five sessions with a total dose of 36.25 Gy, individual sessions with a dose of 7.25 Gy, 3 sessions per week.

### Magnetic Resonance Examinations

MR image acquisition was performed using a 1.5T MRI (Magnetom Avanto, Siemens, Erlangen, Germany). Patients were in the supine position and were examined using a 4-channel body coil placed over the pelvic region. The MRI protocol consisted of axial *T*_2_ weighted, axial diffusion weighted imaging (DWI) and multiecho *T*_2_ weighted images for quantitative *T*_2_ mapping. The imaging parameters are listed in Table 1.

**Table 1:** The parameters of the MR sequences used in this study.

| Sequence Type | FoV<br>(mm) | TE<br>(ms) | TR<br>(ms) | Matrix<br>size | Voxel size<br>(mm) | Flip Angle<br>(°) |
| --- | --- | --- | --- | --- | --- | --- |
| **Axial T2w TSE** | 200 | 89 | 3440 | 256x205 | 1.0x0.8x3.0 | 150 |
| **DWI EPI** | 221 | 92 | 400 | 160x112 | 2.3x1.6x3.0 | 90 |
| **T2maps SE** | 230 | 22, 44, 66, ..., 352 | 3000 | 256x197 | 1.2x0.9x5.0 | 180 |

Note: TSE – turbo spin echo, T2W – T2-weighted, DWI – diffusion weighted imaging, EPI – echo-planar imaging, SE – spin echo, FoV – field of view, TE – echo time (in T2 maps starting echo 22 with step of 22 up to 352 ms), TR – repetition time.

The initial MRI measurement (*M*_0_) was planned prior to commencing the radiation therapy. The procedure was performed after the placement of gold fiducial markers within the prostate for the purpose of aligning the image and providing a guidance throughout the treatment process, ensuring precise targeting, and minimizing potential side effects. The follow-up MRI measurement (*M*_1_) was planned for two weeks after radiation therapy ended. This provided adequate time for the treatment effects to manifest and be accurately evaluated.

After the post-therapy MRI scan, five additional check-ups were arranged (*M*_2_ - *M*_6_), each occurring at monthly intervals to closely monitor any changes or progress. These regular, month-long intervals provided a comprehensive and systematic overview of the patient’s condition, allowing healthcare professionals to track the treatment’s efficacy and adjust their approach as needed. This consistent monitoring was essential to ensuring the most effective possible outcome for the patient. It provided valuable insights into the effectiveness of radiation therapy.

### MR Data Analysis

A comprehensive analysis of the MR images of all 24 patients was conducted under the direction of a radiological expert, P.H., who has more than 20 years of experience in the field. As a first step, the MRI images were loaded into ITK-SNAP, a widely used medical image analysis software program [12], wherein the lesions indicative of prostate cancer (PCa) were meticulously identified and demarcated manually on an initial measurement (*M*_0_) prior to radiation therapy. In order to ensure continuity and consistency of the analysis, the manually created annotations from the initial measurement were replicated on subsequent image sets. These annotations were then manually modified by MR physicists to account for any discernible deformations or displacements in the prostate gland or lesions. Each set of images belonging to each patient was meticulously reviewed and adapted.

Additionally, a trained medical student V.S. manually segmented the entire prostate gland. In accordance with the procedure used for the segmentation of PCa lesions, the segmentation process was rigorously verified by the radiologist, P.H. By implementing this meticulous process, we ensured that the segmentation and annotation process would be accurate and reliable, thereby providing a robust foundation for analysis of the data. The regions of interest (ROIs) identified in each patient were subjected to an in-depth quantitative assessment. The *T*_2_ relaxation times were determined by analyzing *T*_2_-weighted multiecho scans. To provide a comprehensive representation of tissue properties, a total of sixteen echos were incorporated into this study. It is worth noting that the first echo was systematically omitted from the analysis. The removal of the first echo is a common technique in quantitative MRI since it often contains significant system imperfections, allowing for a more robust and reliable fitting of the *T*_2_ relaxation times [13]. The fitting process of *T*_2_ relaxation curve was carried out using an in-house MATLAB (MathWorks, Natick, MA), on a voxel-by-voxel basis. The fitted *T*_2_ relaxation times were then used as the basis for further analysis. By using this approach, we were able to extract detailed and clinically relevant information from the *T*_2_-weighted multi-echo scans allowing better understanding of the radiotherapy impact [14].

### Statistical Analysis

For each of the 24 subjects included in the study, *T*_2_ median values were calculated for the entire prostate. Nevertheless, three subjects were excluded from the statistical analysis as a result of MR artifacts or missed appointments, resulting in a final total of 21 subjects being included. The data were first assessed for normality using the Shapiro-Wilk tests. This test showed that the data were not normally distributed, and hence, a log transformation was applied to correct this. After applying the log transformation [15], data with p-values less than 0.05 were considered to indicate a statistically significant difference. Then, we performed a Repeated Measures Analysis of Variance (RM-ANOVA) to examine the significance of differences in *T*_2_ median values between the seven measurements. This analysis controls subject-level variability and is appropriate for our study design where the same subjects were measured repeatedly. The RM-ANOVA was followed by post-hoc pairwise t-tests to understand where these differences lay. Considering the large number of pairwise comparisons, the Bonferroni correction was applied. A p-value (after Bonferroni correction) less than 0.05 was considered statistically significant.

## Results

Our findings demonstrated a significant association between *T*_2_ relaxation times and an aspect of treatment response. Specifically, The RM-ANOVA demonstrated a statistically significant difference in the *T*_2_ median values across the seven measurements (F (6, 120) = 0.611, p *<<* 0.001, *η*^2^ = 0.305). This suggests that there is a significant effect of measurement time point on *T*_2_ median values. The MRI measurements are denoted as *M*_0_ – *M*_6_, where *M*_0_ is the initial measurement prior the radiotherapy and *M*_1_ - *M*_6_ are the follow-up measurements (*M*_1_ - two weeks after radiotherapy, *M*_2_ - *M*_6_ monthly interval check-ups).

The Bonferroni post hoc tests revealed that *T*_2_ median values were significantly different at baseline (*M*_0_) as well as at all other measurement points (*M*_1_ - *M*_6_). Specifically, a statistically significant difference was found between *M*_0_ and *M*_1_ (T (20) = 4.97, p = 0.0016). This difference continued to increase over time with *T*_2_ median values from *M*_0_ significantly differing from *M*_2_ (T (20) = 10.23, p *<* 0.001), *M*_3_ (T (20) = 12.56, p *<* 0.001), M4 (T (20) = 11.61, p *<* 0.001), M5 (T (20) = 10.49, p *<* 0.001), and M6 (T (20) = 12.33, p *<* 0.001).

Interestingly, a significant difference in *T*_2_ median values was also observed between *M*_1_ and the following time points: *M*_3_ (T (20) = 3.94, p = 0.017), *M*_4_ (T (20) = 3.68, p = 0.031), *M*_5_ (T (20) = 3.53, p = 0.044), and *M*_6_ (T (20) = 3.42, p = 0.057). However, there was no significant difference observed between *M*_1_ and *M*_2_ after Bonferroni correction (T (20) = 3.01, p = 0.146). These results suggest a consistent decrease in *T*_2_ median values from baseline (*M*_0_) through the various stages of radiotherapy, with the exception of *M*_1_ to *M*_2_ where the changes were not statistically significant.

Figure 2 illustrates the variation in *T*_2_ median relaxation times across different measurements. For most patients (denoted using the letters A – X), the most substantial change occurs between baseline (*M*_0_) and the first follow-up measurement (*M*_1_). However, it should be noted that some patients (e.g. patients C, G, and M) exhibit a delayed response, with the greatest change occurring between M0 and the second follow-up (*M*_2_). This emphasizes the inter-individual variability in response to radiotherapy and highlights the importance of longitudinal monitoring to adequately capture this heterogeneity.

**Figure 2:**
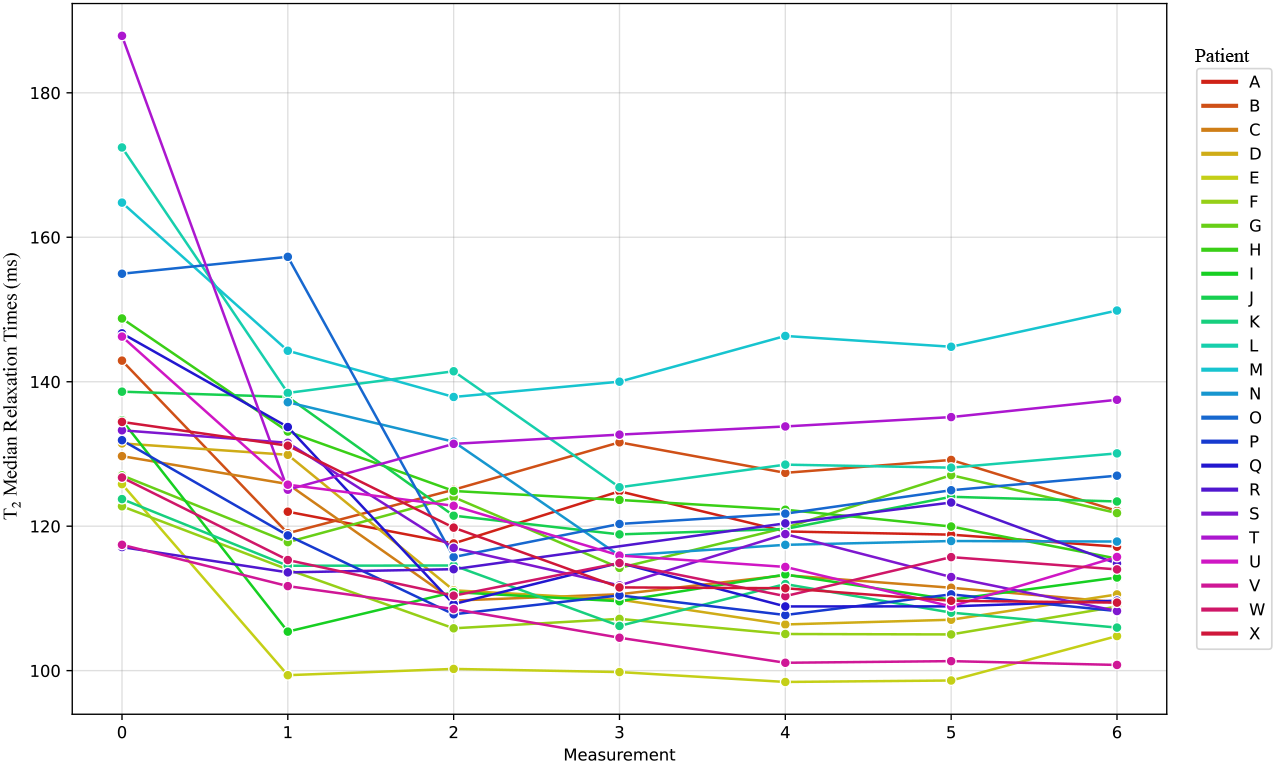
Longitudinal changes in *T*_2_ median relaxation times across all patients (A – X) and measurements. Measurement 0 – initial scan before radiotherapy (*M*_0_). Measurement 1 – first check up two weeks after radiation therapy ended (*M*_1_) and Measurement 2 – 6 monthly intervals check up (*M*_2_ – *M*_6_).

Furthermore, we visualized the changes in *T*_2_ median relaxation time using a heatmap, as shown in Figure 3, where each row corresponds to a specific patient (A – X), and each column represents an individual measurement time point (*M*_0_ – *M*_6_). Each cell in the heatmap is colored according to its median *T*_2_, with darker colors indicating higher values. It provides a comprehensive, color-coded overview of the shifts in *T*_2_ values across the patient cohort and measurement points, allowing for an at-a-glance comparison of therapeutic responses.

**Figure 3:**
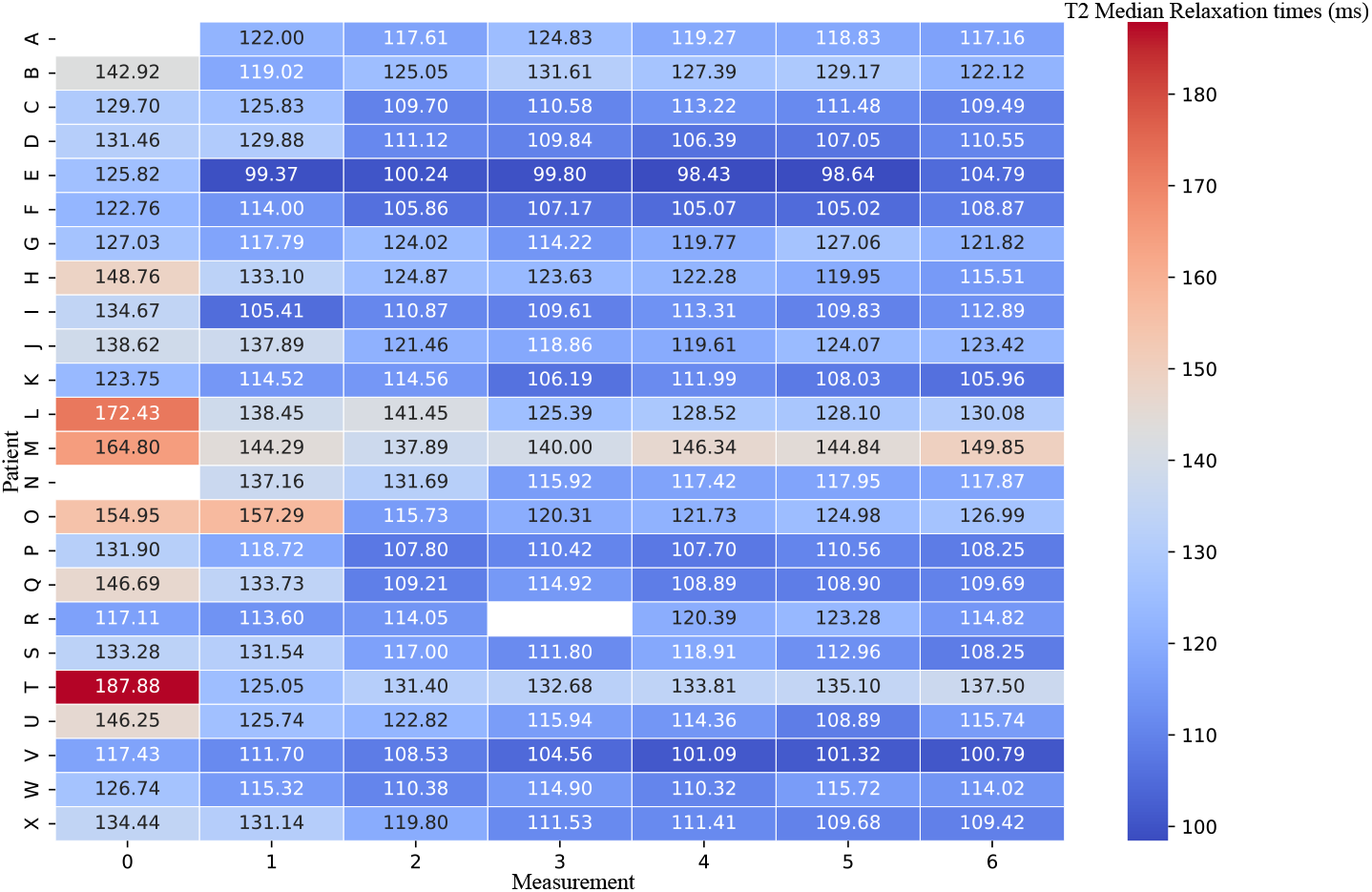
Heatmap of *T*_2_ Median Relaxation Times. Each row represents a patient, and each column represents a distinct measurement time point. Darker colors indicate higher *T*_2_ median values, highlighting the pattern of change over time. Note that the absence of data in some cells is a result of either missed appointments or the occurrence of image artifacts.

Case studies of two patients (Figure 4) offer more detailed insights into these differing therapeutic responses. Patient A exhibited a delayed decrease in *T*_2_ relaxation times, notable in the later measurement time points of the *T*_2_ maps. Despite this initial delay, both diffusion restriction, as seen in the DWI, and a decrease in PSA levels indicate an effective, although prolonged, therapeutic response. Conversely, patient B, who had higher baseline *T*_2_ median values, demonstrated a remarkable therapy response. The 33.44% change in *T*_2_ median values between the baseline (*M*_0_) and first follow-up (*M*_1_) was visually apparent in the *T*_2_ maps and marked one of the most significant responses in our study cohort. This implies that *T*_2_ relaxometry techniques could be a valuable tool in the monitoring of radiotherapy for low-risk prostate cancer patients.

**Figure 4:**
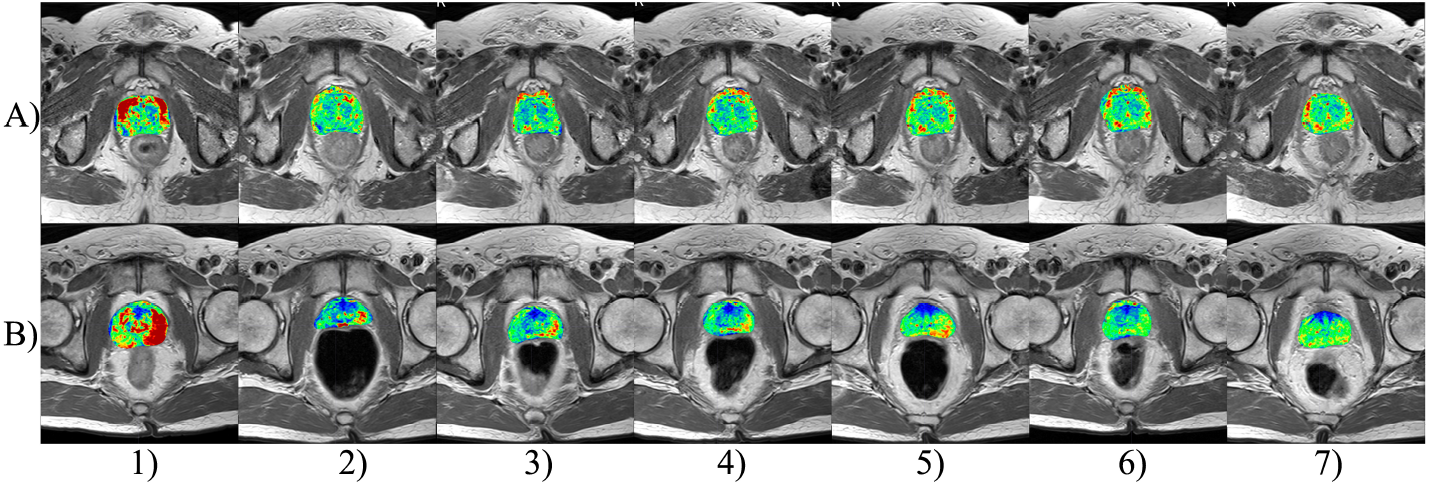
Comparative *T*_2_ maps of two patients with contrasting responses to radiotherapy. Each row represents a distinct measurement time point for each patient, with corresponding *T*_2_ maps overlayed. A) Patient M demonstrates a delayed decrease in *T*_2_ relaxation times, confirmed with weak RT response, visible in later measurement time points. B) Patient T, starting with higher baseline *T*_2_ median values, exhibits a dramatic response to the therapy with a 33.44% change in *T*_2_ median values between baseline (*M*_0_) and the first follow-up (*M*_1_). The *T*_2_ maps visually reflect this marked difference, underscoring the therapeutic impact.

## Discussion

The results of both the statistical analysis and imaging visualizations demonstrate distinct patterns of therapeutic response among patients. As evidenced by the line graph in Figure 2, the most substantial changes in *T*_2_ median relaxation times typically occur between the *M*_0_ and *M*_1_, with the exceptions in some patients where the most significant changes occurred between *M*_0_ and *M*_2_. This is further highlighted using the heatmap of *T*_2_ median values (Figure 3) which provides at-a-glance comparison of therapeutic responses suitable for clinical practice usage.

*T*_2_ relaxometry is a method providing insights into water content of the examined tissue. However, one must keep in mind that a prostate tumor is not the only pathology that can be present in the prostate examined [16]. It can be expected that the subjects involved in this study also suffered from prostatitis: acute or chronic. Acute prostatitis is associated with a higher proportion of water whereas chronic one is linked with higher proportion of fibrosis leading to reduction of *T*_2_ relaxation times [17]. Another factor playing an important role is the proportion of stromal and glandular hyperplasia, which has more water than hypertrophic stroma [18]. These proportions are variable and cannot be generalized. Therefore, it can be expected that the output for each individual (its range and baseline) will differ as shown in our results.

Another aspect that could affect the results of the start scan was the fact that it was always measured after the implantation of gold fiducials into the prostate, used for navigation of subsequent radiotherapy using the CyberKnife. This application may have been a cause of acute prostatitis in the given patient. In addition, prostatitis is also activated after radiotherapy itself, when some patients develop so-called post-radiation prostatitis, leading to PSA elevations and decreases in the period up to several weeks after radiotherapy.

The prostatitis and its share in the stromal and acinar part greatly changes the resulting relaxation times. For such a limited cohort, it is not possible to correlate individual times, but it is possible to compare their trend, as showed by Foltz et al. [19]. In Foltz’s study, the measurements were carried bi-weekly throughout eight weeks of radiotherapy and the results proved that *T*_2_ can serve as a useful biomarker to detect early response to radiotherapy even in patients with low and intermediate risk localized prostate cancer.

A delayed decrease in relaxation times is evident in one patient (M). In this case, a long-term restriction in diffusion was demonstrated in this patient when examined using the DWI, which indicates that the effect of the therapy was prolonged. However, there was a decrease in PSA, which correlates with bio-chemical markers of therapy effectiveness according to oncological procedures, so this case was not evaluated as a therapy failure.

Finally, the study presented herein offered some valuable insights but also faced several limitations, for example, the absence of a control group or reference from histopathological evaluation. The main reason for not obtaining a detailed histopathological evaluation and a deeper specification of the tumor tissue distribution was not performed, because the radical surgical procedure was found unnecessary in these patients due to low grade staging. Therefore, the entire prostate volume was not processed. Because of low staging, only a needle biopsy was performed and radiotherapy was initiated. As a result of the collection of biopsy samples, it is not possible to describe in detail the distribution and specification of tumor cells within the tumor mass, but only to provide a rough estimate of the patient’s prognosis based on a risk stratification. The reason for this is that samples are taken from different parts of the prostate, which (despite the large number of samples taken) does not cover the entire organ and does not represent the tumor tissue, as it would if the entire prostate was removed [20, 21].

Furthermore, instead of selecting the ROI (the tumor) whole prostate was analyzed, which may lead to bias due to other mechanisms and pathologies taking place in the affected organ. However, since the focus of the study was on low-grade tumors, the determination of the ROI is complicated and hardly reproducible for repeated measurements. Additionally, the use of an endorectal coil could significantly enhance the image quality and allow magnetic resonance spectroscopy, however, because of the many repeated measurements, its use has been prohibited.

## Conclusion

In this study, we used 16 echo times to determine the *T*_2_. This type of calculation is considered very precise. Moreover, the cohort selected consisted of 24 elderly patients with low grade carcinoma, which can be considered as unique cohort. The patients were scanned 7 times, once before radiotherapy and then monthly, which provides more reasonable window to monitor the radiotherapy effect. The main contributions can be summarized as follows:

1. Quantification of scarring – description of the scarring process in the tumor in well-responsive tumors.
2. Recognition of therapy failure – the results of this study showed a demonstrable reduction in the relaxation times in a patient with a prolonged lesion. It is especially important during the period of post-radiation prostatitis, when extreme PSA elevation does not mean therapy failure.
3. Radiotherapy response monitoring in unconfirmed patients – in the cases when a less risky part of the tumor was removed during the prostate biopsy, the determined Gleason score does not correspond to the real risks and prescribed treatment (e.g. cases of misdiagnosed multifocal carcinoma [22]). *T*_2_ relaxometry is a promising tool to monitor the response to radiotherapy even in unconfirmed, but still at-risk patients for moderate or high-risk cancers (Gleason score above 4 + 3 to 5 + 5).

The changes in *T*_2_ values during the course of radiotherapy can help in uncovering alterations in cancer and prostate tissue more sensitively than other parameters, if monitored and visualized appropriately.

## Data Availability

All data produced in the present study are available upon reasonable request to the authors

## Acknowledgment

This work was supported by Ministry of Health, Czech Republic - conceptual development of research organization (FNOs/2020) and by the Ministry of Education of Czech Republic under Project SP2023/042.

## References

[1] A. K. Miyahira, A. Sharp, L. Ellis, J. Jones, S. Kaochar, H. B. Larman, D. A. Quigley, H. Ye, J. W. Simons, K. J. Pienta, H. R. Soule, Prostate cancer research: The next generation; report from the 2019 Coffey-Holden Prostate Cancer Academy Meeting, The Prostate 80 (2) (2020) 113–132. doi:10.1002/pros.23934.

[2] C. Ritch, M. Cookson, Recent trends in the management of advanced prostate cancer, F1000Research 7 (2018) 1513. doi:10.12688/f1000research.15382.1.

[3] Z. Khan, N. Yahya, K. Alsaih, M. I. Al-Hiyali, F. Meriaudeau, Recent Automatic Segmentation Algorithms of MRI Prostate Regions: A Review, IEEE Access 9 (2021) 97878–97905. doi:10.1109/ACCESS.2021.3090825.

[4] G. Litjens, O. Debats, J. Barentsz, N. Karssemeijer, H. Huisman, Computer-Aided Detection of Prostate Cancer in MRI, IEEE Transactions on Medical Imaging 33 (5) (2014) 1083–1092. doi:10.1109/TMI.2014.2303821.

[5] G. Lemaitre, R. Marti, J. Freixenet, J. C. Vilanova, P. M. Walker, F. Meriaudeau, Computer-Aided Detection and diagnosis for prostate cancer based on mono and multi-parametric MRI: A review, Computers in Biology and Medicine 60 (2015) 8–31. doi:10.1016/j.compbiomed.2015.02.009.

[6] B. Turkbey, A. B. Rosenkrantz, M. A. Haider, A. R. Padhani, G. Villeirs, K. J. Macura, C. M. Tempany, P. L. Choyke, F. Cornud, D. J. Margolis, H. C. Thoeny, S. Verma, J. Barentsz, J. C. Weinreb, Prostate Imaging Reporting and Data System Version 2.1: 2019 Update of Prostate Imaging Reporting and Data System Version 2, European Urology 76 (3) (2019) 340–351. doi:10.1016/j.eururo.2019.02.033.

[7] A. Panda, V. Gulani, Quantitative Imaging of Prostate: Scope and Future Directions, in: A. Panda, V. Gulani, L. Ponsky (Eds.), Reading MRI of the Prostate, Springer International Publishing, Cham, 2020, pp. 97–108. doi:10.1007/978-3-319-99357-7_10.

[8] S. C. Deoni, Quantitative Relaxometry of the Brain, Topics in Magnetic Resonance Imaging 21 (2) (2010) 101–113. doi:10.1097/RMR.0b013e31821e56d8.

[9] A. A. O. Carneiro, G. R. Vilela, D. B. D. Araujo, O. Baffa, MRI relaxometry: Methods and applications, Brazilian Journal of Physics 36 (1a) (Mar. 2006). doi:10.1590/S0103-97332006000100005.

[10] S. Schneider, R. I. Jølck, E. G. C. Troost, A. L. Hoffmann, Quantification of MRI visibility and artifacts at 3T of liquid fiducial marker in a pancreas tissue-mimicking phantom, Medical Physics 45 (1) (2018) 37–47. doi: 10.1002/mp.12670.

[11] L. Knybel, J. Cvek, T. Blazek, A. Binarova, T. Parackova, K. Resova, Prostate deformation during hypofractionated radiotherapy: An analysis of implanted fiducial marker displacement, Radiation Oncology 16 (1) (2021) 235. doi:10.1186/s13014-021-01958-4.

[12] P. A. Yushkevich, Y. Gao, G. Gerig, ITK-SNAP: An interactive tool for semi-automatic segmentation of multi-modality biomedical images, in: 2016 38th Annual International Conference of the IEEE Engineering in Medicine and Biology Society (EMBC), IEEE, Orlando, FL, USA, 2016, pp. 3342–3345. doi:10.1109/EMBC.2016.7591443.

[13] D. Milford, N. Rosbach, M. Bendszus, S. Heiland, Mono-Exponential Fitting in T2-Relaxometry: Relevance of Offset and First Echo, PLOS ONE 10 (12) (2015) e0145255. doi:10.1371/journal.pone.0145255.

[14] A. Chatterjee, A. Devaraj, M. Mathew, T. Szasz, T. Antic, G. S. Karczmar, A. Oto, Performance of T2 Maps in the Detection of Prostate Cancer, Academic Radiology 26 (1) (2019) 15–21. doi:10.1016/j.acra.2018.04.005.

[15] J. Osborne, Notes on the use of data transformations, Practical Assessment, Research, and Evaluation 8 (6). doi:10.7275/4VNG-5608.

[16] A. Stabile, F. Giganti, A. B. Rosenkrantz, S. S. Taneja, G. Villeirs, S. Gill, C. Allen, M. Emberton, C. M. Moore, V. Kasivisvanathan, Multiparametric MRI for prostate cancer diagnosis: Current status and future directions, Nature Reviews Urology 17 (1) (2020) 41–61. doi: 10.1038/s41585-019-0212-4.

[17] A. B. Rosenkrantz, S. S. Taneja, Radiologist, Be Aware: Ten Pitfalls That Confound the Interpretation of Multiparametric Prostate MRI, American Journal of Roentgenology 202 (1) (2014) 109–120. doi:10.2214/AJR.13.10699.

[18] J. C. Weinreb, J. O. Barentsz, P. L. Choyke, F. Cornud, M. A. Haider, J. Macura, D. Margolis, M. D. Schnall, F. Shtern, C. M. Tempany, H. C. Thoeny, S. Verma, PI-RADS Prostate Imaging – Reporting and Data System: 2015, Version 2, European Urology 69 (1) (2016) 16–40. doi:10.1016/j.eururo.2015.08.052.

[19] W. D. Foltz, A. Wu, P. Chung, C. Catton, A. Bayley, M. Milosevic, R. Bristow, P. Warde, A. Simeonov, D. A. Jaffray, M. A. Haider, C. Menard, Changes in apparent diffusion coefficient and T _2_ relaxation during radiotherapy for prostate cancer, Journal of Magnetic Resonance Imaging 37 (4) (2013) 909–916. doi:10.1002/jmri.23885.

[20] D. G. Bostwick, Evaluating prostate needle biopsy: Therapeutic and prognostic importance, CA: A Cancer Journal for Clinicians 47 (5) (1997) 297–319. doi:10.3322/canjclin.47.5.297.

[21] E. Short, A. Y. Warren, M. Varma, Gleason grading of prostate cancer: A pragmatic approach, Diagnostic Histopathology 25 (10) (2019) 371–378. doi:10.1016/j.mpdhp.2019.07.001.

[22] A. Ali, A. Du Feu, P. Oliveira, A. Choudhury, R. G. Bristow, E. Baena, Prostate zones and cancer: Lost in transition?, Nature Reviews Urology 19 (2) (2022) 101–115. doi:10.1038/s41585-021-00524-7.

